# Diagnostic Criteria and Disease Staging for Desmoplakin Cardiomyopathy

**DOI:** 10.1101/2025.06.16.25329734

**Authors:** Eric Smith, Alessio Gasperetti, Richard T Carrick, Alexandros Protonotarios, Petros Syrris, Barbara Bauce, Kalliopi Pilichou, Brittney Murray, Crystal Tichnell, Cristina Basso, Paul Anders Sletten Olsen, Chiara Cappelletto, Victoria N. Parikh, Liang Chen, Stacey Peters, Antonella Mancinelli, Anna Maria Iorio, Roberta Scotto, Julia Cadrin-Tourigny, Nisha A. Gilotra, Manuela Iseppi, Giovanni Peretto, Marco Schiavone, Mark Abela, Daniela Vargas, Cinzia Crescenzi, Leonardo Calò, James Ware, Lia Crotti, Michele Ciabatti, Michela Casella, Alexandra Apostu, Ruxandra Jurcut, Claudia Raineri, Filippo Angelini, Carlos Fernández-Sellers, Esther Zorio, Sing-Chien Yap, Moniek G Cox, Arman Salavati, Anneline te Riele, Arthur Wilde, Ahmad S. Amin, Peter van Tintelen, Ardan M. Saguner, Firat Duru, Dominic Abrams, Marina Cerrone, Maddalena Graziosi, Elena Biagini, Juan Ramon Gimeno, Estelle Gandjbakhch, Neal Lakdawala, Maurizio Pieroni, Marco Merlo, Gianfranco Sinagra, Kristina Haugaa, Elena Arbelo, Perry M. Elliott, Matthew Taylor, Luisa Mestroni, Hugh Calkins, Cynthia A. James, Adam S. Helms

## Abstract

**Background:** Desmoplakin (DSP) cardiomyopathy, caused by variants in the gene *DSP*, is a unique subtype of cardiomyopathy distinct from typical dilated or arrhythmogenic right ventricular cardiomyopathies. Specific diagnostic and disease staging criteria have yet to be developed for DSP cardiomyopathy.

**Objective:** Utilizing a large cohort of DSP cardiomyopathy patients and their genotype-positive family members, this study aims to develop diagnostic and disease staging criteria for DSP cardiomyopathy.

**Methods:** Patients from the *DSP*-ERADOS Network with complete rhythm monitoring, electrocardiogram and cardiac magnetic resonance imaging were enrolled. Diagnostic criteria were assessed in initially-presenting patients (probands) and their genotype-positive family members. Early disease criteria (with preserved left ventricular ejection fraction, LVEF) were integrated into standard LVEF-based classifications. Diagnostic and staging criteria were assessed by time-event analyses (major ventricular arrhythmia and heart failure events).

**Results:** A total of 605 patients with complete diagnostic testing were included (mean age 40 yr, 60% female, 40% probands). The most prevalent disease features in probands were premature ventricular contractions (PVCs) >500/24hr (66%), nonsustained ventricular tachycardia (NSVT, 29%), LV late gadolinium enhancement (LGE, 53%), and reduced LVEF (44%). The presence of any one of these features was 97% sensitive for diagnosis (along with a *DSP* pathogenic variant) and were therefore considered as diagnostic criteria. Using these criteria, 77% of genotype-positive family members were considered clinically affected. Isolated right ventricular (RV) involvement occurred in only 0.7%. The absence of diagnostic criteria identified a low-risk group (composite event rate 0.8%/year, p<0.001). Integration of these criteria into LVEF-based classification improved identification of composite arrhythmia/heart failure events (early: diagnostic criteria with LVEF >50%, HR 2.7, p=0.04; intermediate: LVEF 41-49%, HR 3.7, p=0.009; advanced: LVEF <40% HR 10.3, p<0.001). LGE was mostly subepicardial (87%). Circumferential (ring-like) LGE was more frequent in intermediate or advanced vs early disease (66% vs 48%, p<0.001).

**Conclusion:** This study identifies genotype-specific diagnostic and disease staging criteria for DSP cardiomyopathy that improve identification of risk for both heart failure and sustained ventricular arrhythmias. This work highlights how gene-specific criteria may be used to refine diagnosis and staging for cardiomyopathy subtypes – a critical step as gene-targeted treatments move toward clinical trials.

## Introduction

The identification of genetic etiologies has transformed our understanding of cardiomyopathy. Disease characteristics have been shown to strongly stratify by genotype, suggesting that traditional descriptive categories (e.g. “dilated cardiomyopathy”) inadequately capture the defining features of many inherited cardiac diagnoses. Thus, a reliance on diagnostic criteria developed for broadly inclusive disease categories may hinder accurate diagnosis of specific genetic subtypes. Moreover, natural history and risk burden has been shown to be highly dependent on genetic etiology, as has now been demonstrated for several cardiomyopathy-associated genes, including desmoplakin (*DSP)*.^1–6^

Desmoplakin (DSP) cardiomyopathy, caused by variants in *DSP*, is a subtype of cardiomyopathy distinct from either typical dilated cardiomyopathy (DCM) or arrhythmogenic right ventricular cardiomyopathy (ARVC), but with overlapping features of both.^1, 7–9^ DSP cardiomyopathy is one of the most common genetic subtypes of all DCM/ARVC combined.^5, 10, 11^ Standard diagnostic criteria developed for either DCM or ARVC have been shown to poorly identify DSP cardiomyopathy.^1, 12, 13^ Consensus criteria developed for a separate category of “arrhythmogenic left ventricular cardiomyopathy” (ALVC) may better apply for DSP cardiomyopathy but the individual criteria have not been tested in a deeply phenotyped cohort of patients with *DSP* variants.^14^ Meanwhile, heart failure guidelines classify progression of DCM primarily based on left ventricular ejection fraction. No systematic studies have tested diagnostic or disease staging criteria for DSP cardiomyopathy. Given the increased availability of broad genetic testing and recent emergence of gene-specific targeted treatment approaches, a gene-specific diagnostic and clinical staging approach will be important for prognosis, management, and genotype-specific clinical trials of DSP cardiomyopathy.

A major hurdle to establishing both diagnostic and gene-specific disease staging criteria has been a lack of large cohorts with comprehensive rhythm monitoring and cardiac magnetic resonance imaging (MRI). Extended rhythm monitoring and cardiac MRI are both essential for identification of early signs of DSP cardiomyopathy but have been incompletely captured in prior cohorts.^1, 7–9^ To address this limitation, we assembled a large international cohort comprised solely of individuals who had completed comprehensive baseline testing, including both cardiac MRI and extended rhythm monitoring. Importantly, the cohort consisted of both initially presenting patients (probands) with DSP cardiomyopathy and their genetic test positive relatives to ensure inclusion of a broad spectrum of disease severity, including many individuals with preserved left ventricular ejection fraction (LVEF). Using this approach, we define diagnostic criteria for DSP cardiomyopathy and demonstrate the importance of these criteria, in particular, for early-stage disease. Additionally, we integrated these criteria into the standard heart failure LVEF-based classification of current ESC and AHA/ACC guidelines.^15, 16^ This staging scheme resulted in more precise risk stratification for both heart failure and composite arrhythmia outcomes compared to LVEF-only based classification (**Figure 1, Central Figure**). Our approach for DSP cardiomyopathy highlights how gene-specific criteria may be used to refine diagnosis and staging for cardiomyopathy subtypes – a critical step as gene-targeted treatments advance toward clinical trials.

**Figure 1.**
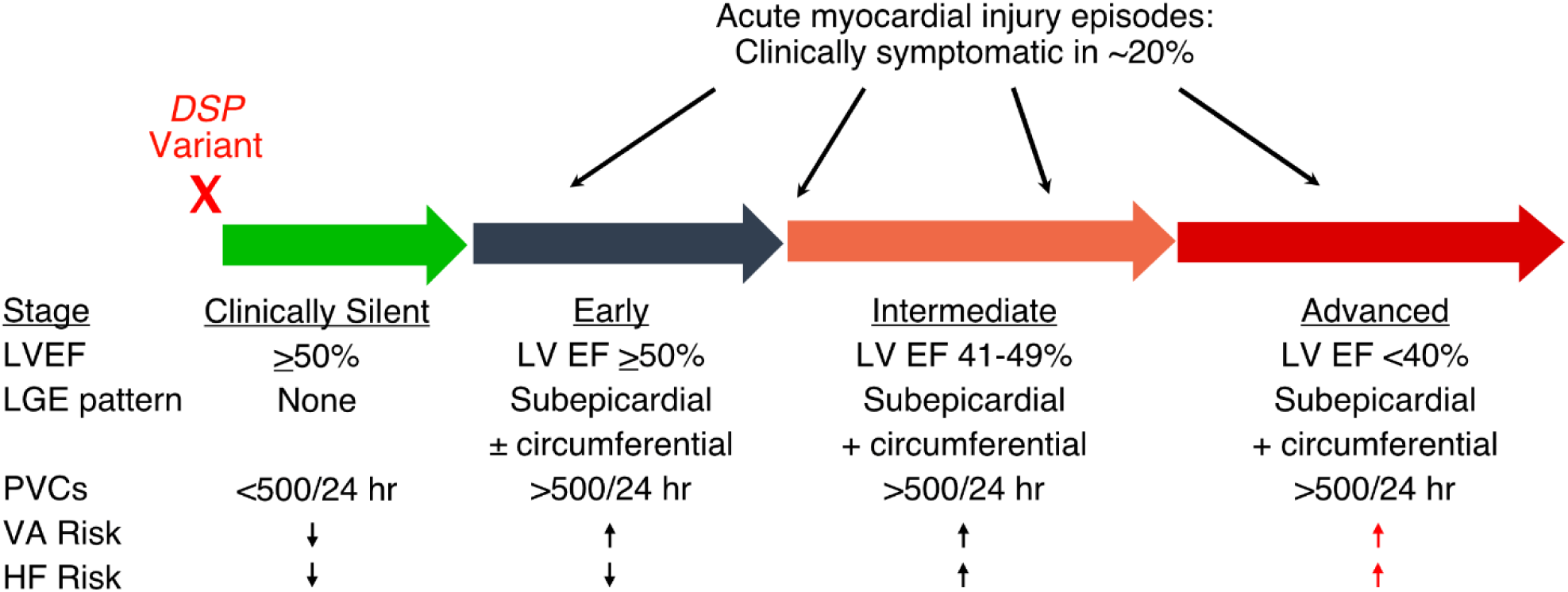
(Central Figure). Proposed disease stages for DSP cardiomyopathy.

## METHODS

### Study population

Patients enrolled in the *DSP*-ERADOS (Desmoplakin SPecific Effort for a RAre Disease Outcome Study) Network were used for this study (N=1464 at the time of the data extraction). All individuals in the DSP-ERADOS cohort have a confirmed pathogenic or likely pathogenic (P/LP) variant. Only individuals with complete rhythm monitoring, ECG, and cardiac MRI with comprehensive LGE segmentation were included for the present study (N=605). As previously described, the *DSP*-ERADOS Network is an international collaboration currently including 43 institutions across 14 countries (United States, Canada, Italy, United Kingdom, the Netherlands, France, Germany, Spain, Australia, China, Malta, Switzerland, Norway and Romania), with each institution functioning as an independent, prospective, observational patient registry.^17^

### Data Collection

The data collection strategy for patients enrolled in the *DSP*-ERADOS Network has been described before.^9^ In brief, demographics, patient medical history, genetic test results, and cardiac diagnostic testing (12-lead electrocardiogram (ECG), cardiac magnetic resonance (CMR), 24 h Holter ECG monitoring) were retrieved for each patient by each involved institution at the time of first clinical assessment (defined as patient baseline). Outcomes were adjudicated locally at each participating center via review of patient medical records, including implantable cardioverter-defibrillator (ICD) interrogations. Probands were defined as the first affected individual in a family who came to medical attention. All *DSP* genetic variants categorized as P or LP locally underwent centralized expert review by specialists in cardiac genetics (B.M., C.A.J.), in accordance with the American College of Medical Genetics and Genomics (ACMG) guidelines and previously published ACM specific ACMG adjustments.^18, 19^

### Adverse Event Definitions

Overall composite adverse events were defined as either a major arrhythmia or major heart failure event. Major arrhythmia events consisted of sustained ventricular arrhythmia, appropriate ICD intervention, ventricular fibrillation (VF), or sudden cardiac arrest. Major heart failure events consisted of heart failure hospitalization, heart transplant or left ventricular assist device placement.

### MRI Segmentation

A 36-segment MRI model was used to capture regional segments (septal, anterior, lateral, inferior), ventricular level (base, middle, or apex), and muscle layer (subepicardial, midmyocardial, or subendocardial). LGE presence was determined as a binary variable (present or absent) for each of these 36 segments.

### Statistical Analysis

All analysis were performed using STATA v14.0 (StataCorp, 4905 Lakeway Dr, College Station, TX, USA). Nominal variables were expressed as numbers (%), and continuous variables as mean±standard deviation or median (interquartile range [IQR]), as appropriate according to distribution. Comparisons for binary variables were performed by Chi-square or Fisher’s exact tests. For continuous variables, independent t-tests or Mann-Whitney U tests were used. Time to event analysis was performed through a Cox regression analysis, using putative diagnostic criteria alone or in combinations as variables. These analyses were also performed with 1) limiting the cohort to preserved LV EF (defined as LVEF ≥50%) and 2) stratifying the cohort by putative disease stages. Stages of disease were as defined in the manuscript (silent; early; intermediate; advanced). LV EF categories were defined in alignment with both the ACC/AHA and ESC heart failure guidelines (i.e., ≥50%, 41-49%, and ≤40%).^15, 16^ Time zero was set at the time of first clinical assessment. Censoring was set at last available follow up, occurrence of any of the events after first clinical assessment, or at 5 years (a 5 year limit was used to minimize effects of interval disease progression following the baseline evaluation). Strength of association of predictors with events was reported using hazard ratios (HR). Survival from occurrence of events was graphically displayed using Kaplan Meier curves stratified according to putative disease stages. A p value < 0.05 was considered statistically significant.

## Results

### Baseline Characteristics of DSP Cardiomyopathy Patients and Their Genotype-Positive Family Members

The cohort for the present study consisted of 605 individuals who had complete baseline data for resting electrocardiogram (ECG), ambulatory rhythm monitoring, and cardiac MRI. The average age of the cohort was 40.4 ± 16.5 yr and 60% were female. P/LP *DSP* variants were primarily truncating (84%). The cohort was comprised of 241 probands and 364 genotype-positive family members. Since probands often have more severe disease than their genotype-positive family members, we examined baseline clinical variables stratified by proband status (**Table 1**). PVC count was higher in probands (1,125 vs 243 PVCs/day, p<0.001). When compared using various thresholds, probands consistently had frequent PVCs more often than family members – though a notably high proportion of family members also exhibited frequent PVCs across cut-points (e.g. 42% with more than 500 PVCs per 24 hours, **Table 1**). Non-sustained ventricular tachycardia (NSVT) was less common than frequent PVCs but still occurred in a significant proportion of patients (36% for probands vs 12% for family members, p<0.001). Notably, some patients with NSVT did not have frequent PVCs (supplemental table 1). Since both NSVT and frequent PVCs were independently associated with VA risk in DSP cardiomyopathy previously^17^, these characteristics were considered as a combined diagnostic criterion going forward, with PVCs>500/24hr used as the PVC threshold. MRI analysis demonstrated that a high percentage of both probands and family members exhibited LGE (81% for probands vs 59% for family members, p<0.001). When present, LGE was most often subepicardial in both probands and family members (176 of 196, or 90%, for probands with LGE and 178 of 213, or 84%, for family members with LGE). A substantial proportion of individuals were observed to have a reduced LVEF, which was similarly more prevalent in probands (LVEF <50% present in 53% of probands vs 21% of family members, p <0.001). T wave inversions (TWIs) were present in both groups, though the overall prevalence was lower than the above characteristics (**Table 1**). A small proportion in each group was observed to have right ventricular systolic dysfunction (18% vs 6%, p<0.001). In summary, this comprehensive diagnostic evaluation demonstrated that the most common clinical findings in patients with P/LP *DSP* variants are 1) frequent PVCs/NSVT, 2) LGE (most often subepicardial), and 3) reduced LVEF.

**Table 1:** Baseline Characteristics. PVC: premature ventricular contraction, NSVT: non-sustained ventricular tachycardia, MRI: magnetic resonance imaging, LGE: late gadolinium enhancement LVEF: left ventricular ejection fraction, RVEF: right ventricular ejection fraction

| Variable | Probands<br>(n=241) | Non-Probands<br>(n=364) | p-value |
| --- | --- | --- | --- |
| Age (years) | 41.2 ±15.4 | 39.8 ± 17.3 | 0.325 |
| Female Sex | 148 (61.4) | 215 (59.1) | 0.564 |
| European ancestry | 224 (92.9) | 351 (96.4) | 0.053 |
| Truncating variant | 194 (80.5) | 315 (86.5) | 0.047 |
| PVC, median | 1253<br>[300 - 3763] | 243<br>[6 - 1561] | <0.001 |
| PVCs ≥250/24hr | 184 (76.4) | 181 (49.7) | <0.001 |
| PVCs ≥500/24hr | 166 (68.9) | 152 (41.8) | <0.001 |
| PVCs ≥1000/24hr | 138 (57.3) | 116 (31.9) | <0.001 |
| NSVT | 87 (36.1) | 45 (12.4) | <0.001 |
| NSVT or PVC ≥250/24hr | 204 (84.7) | 186 (51.1) | <0.001 |
| NSVT or PVC >500/24hr | 190 (78.8) | 160 (44.0) | <0.001 |
| NSVT or PVC >1000/24hr | 167 (69.3) | 129 (35.4) | <0.001 |
| T-wave inversion in 2 precordial leads | 97 (40.3) | 66 (18.1) | <0.001 |
| T-wave inversion in >3 precordial leads | 76 (31.5) | 42 (11.5) | <0.001 |
| T-wave inversion in inferolateral leads (at least 2 of I, aVL, V5, V6) | 87 (36.1) | 42 (11.5) | <0.001 |
| MRI LGE | 196 (81.3) | 213 (58.5) | <0.001 |
| MRI Subepicardial LGE | 176 (73.0) | 178 (48.9) | <0.001 |
| LVEF <55% | 176 (73.0) | 144 (39.6) | <0.001 |
| LVEF <50% | 128 (53.1) | 77 (21.2) | <0.001 |
| RV systolic dysfunction (RVEF <45%) | 44 (18.3) | 23 (6.3) | <0.001 |

We next assessed the extent to which combinations of these 3 primary characteristics are overlapping by calculating intersections and unions (Supplemental Table 1). As shown by the Venn diagram (**Figure 2A**), incomplete overlap was present among patients with these diagnostic features. While LGE and PVCs/NSVT were often present together (21.7%), LGE and frequent PVCs/NSVT each occurred as isolated findings in substantial proportions (45.9% and 30.9%, respectively), indicating that these variables independently contribute to diagnosis. Notably, most patients with LVEF<50% exhibited both LGE and frequent PVCs/NSVT (**Figure 2A**). Taken together, these diagnostic criteria (**Table 2**) were 97% sensitive for identifying DSP cardiomyopathy probands. Additionally, using this combination of diagnostic features, cardiomyopathy penetrance in genotype-positive family members was 77% at a mean age of 37±17 years.

**Figure 2.**
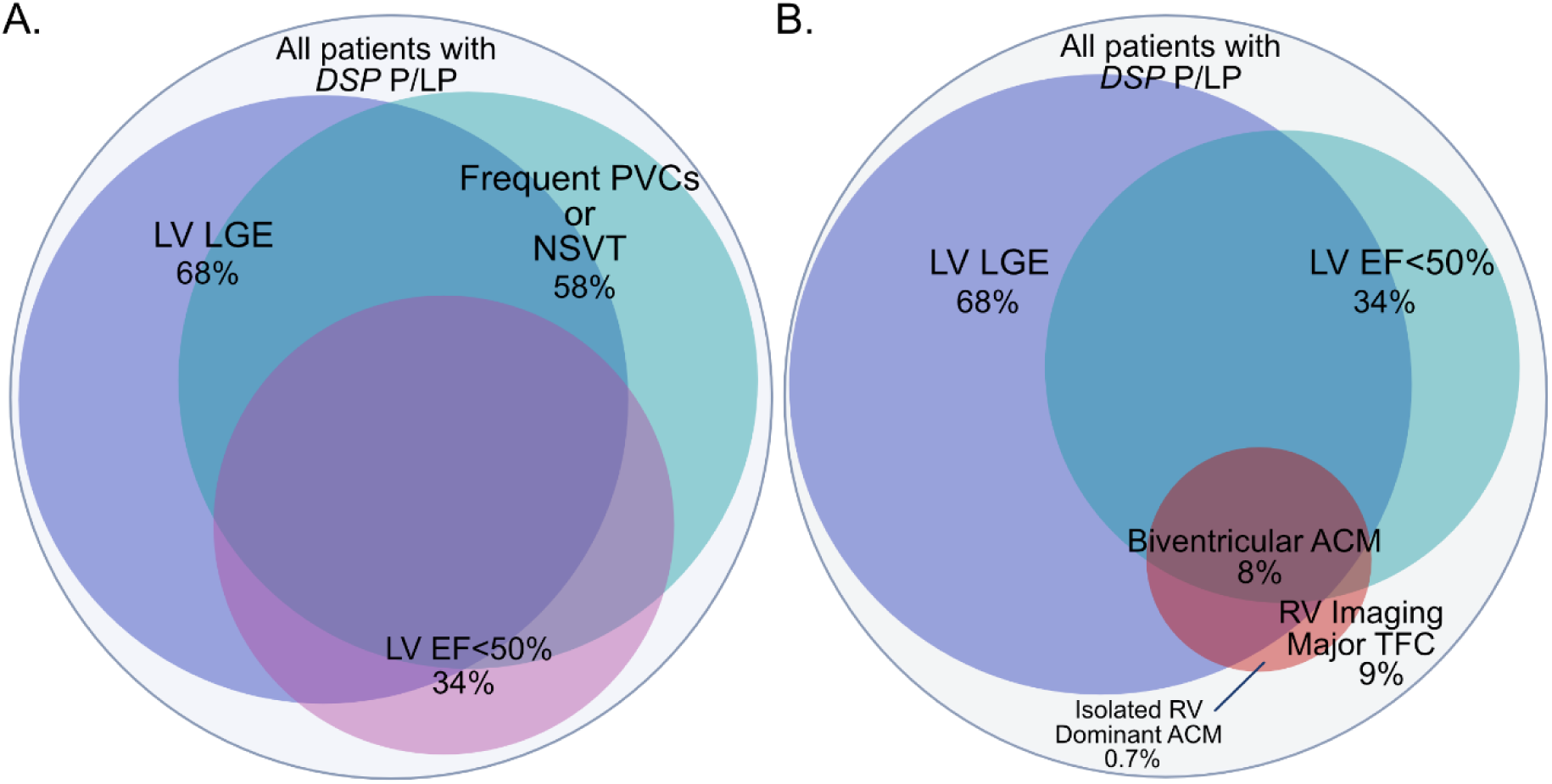
Diagnostic Feature Overlap in DSP Cardiomyopathy. **A.** LV LGE, frequent PVCs or NSVT, and LVEF <50% are prevalent features of DSP cardiomyopathy with substantial but incomplete overlap shown by scaled Venn diagram. **B.** Left ventricular involvement (indicated by LV LGE and/or LVEF <50%) is most common in DSP cardiomyopathy while a small proportion exhibit biventricular involvement and rare patients may exhibit isolated RV involvement, shown by scaled Venn diagram.

**Table 2:** Diagnostic Criteria for DSP Cardiomyopathy. PVC: premature ventricular contraction, NSVT: non-sustained ventricular tachycardia, LV: left ventricle, LGE: late gadolinium enhancement, EF: ejection fraction

|  |
| --- |
| **DSP P/LP Variant, plus any of the following:** |
| Frequent PVCs (>500/24hr) or NSVT |
| Left ventricular LGE (most often subepicardial) |
| LV EF<50% |

### Left and Right Ventricular Involvement

Investigators in our group and others have previously reported that typical ARVC presentations are rare in DSP cardiomyopathy.^1, 8, 9, 20^ However, a remaining question has been whether atypical cases could present with more ARVC features that would require a combined diagnostic approach. Therefore, leveraging this cohort with complete MRI data, we also assessed for the presence of right ventricular structural abnormalities using the 2010 ARVC task force criteria (TFC) for MRI findings in ARVC. Right ventricle (RV) structural involvement was present in only 8.8% of the cohort, 8.1% of whom had biventricular involvement (**Figure 2B**, Supplemental Table 1). In contrast, left ventricle (LV) involvement was present in 74.1% (**Figure 2B**, Supplemental Table 1). Only 4 patients (0.7%) were reported to have isolated RV structural involvement. Thus, RV-specific MRI criteria from the TFC did not yield significant incremental diagnostic yield for DSP cardiomyopathy.

### Adverse Events Based on Presence of Diagnostic Criteria

To verify the clinical significance of the above three primary diagnostic features, we assessed the incidence of composite adverse events over a five-year period using a multivariable Cox regression model. When considered as univariate predictors frequent PVCs/NSVT (p<0.001), and LVEF <50% (p<0.001) were each associated with adverse events (**Table 3**). When considered in the multivariable model, frequent PVCs/NSVT (p<0.002), and LVEF <50% (p=0.001) remained predictors of adverse events (**Table 3**).

**Table 3:** Multivariable Cox Regression Analysis for Composite Adverse Events, Stratified by Diagnostic Criteria. Composite adverse included both major ventricular arrhythmia (sustained ventricular arrhythmia, appropriate ICD intervention, VF, or sudden cardiac arrest) and major heart failure events (heart failure hospitalization, heart transplant, or left ventricular assist device placement). PVC: premature ventricular contraction, NSVT: non-sustained ventricular tachycardia, LV: left ventricle, LGE: late gadolinium enhancement, EF: ejection fraction, HR: hazard ratio, C.I.: Confidence interval

| Variable | Univariate |  |  | Multivariable |  |  |
| --- | --- | --- | --- | --- | --- | --- |
|  | HR | C.I. | p-value | HR | C.I. | p-value |
| PVCs 500+/24hr or NSVT | 3.36 | 1.96–5.76 | <0.001 | 2.52 | 1.40–4.53 | 0.002 |
| LV LGE | 1.39 | 0.87–2.23 | 0.166 | 0.91 | 0.56–1.48 | 0.717 |
| LV EF<50% | 2.82 | 1.87–4.24 | <0.001 | 2.08 | 1.34–3.24 | 0.001 |

We next tested whether frequent PVCs/NSVT and/or LGE were associated with adverse events in patients with a preserved LVEF, since this subset of patients was common in the cohort (46% of the cohort). In patients with LVEF ≥ 50%, no significant difference was identified for composite adverse events in the presence of either frequent PVCs/NSVT or LGE in isolation. However, the presence of both frequent PVCs/NSVT and LGE together was associated with significantly increased risk for the combined outcome compared to individuals with neither of these findings (HR 4.77, p<0.001, Supplemental Figure 1).

Taken together, this analysis demonstrates that frequent PVCs/NSVT and LVEF<50% are independently associated with major heart failure and major ventricular arrhythmia events. Additionally, in patients with LVEF ≥50%, the presence of LGE contributes to risk in combination with PVCs/NSVT.

### Disease Staging for DSP Cardiomyopathy

We next sought to establish disease stages for DSP cardiomyopathy that would integrate with existing heart failure guidelines-based classification. First, we tested whether existing heart failure classifications based on LVEF (i.e., ≥50%, 41%-49%, and ≤40%) identify different levels of risk based on composite events that include major heart failure events. As shown in Supplemental Figure 2, stratification by LVEF did result in a stepwise increase in composite events. We performed a multivariable analysis to test both LVEF classifications and extent of LGE for association with combined heart failure and ventricular arrhythmias. This analysis confirmed that the lowest LVEF category (LVEF<40%) was independently associated with increased composite events (HR 4.58, p<0.001, Supplemental Table 2). However, our analyses above had revealed that early-stage disease features are present with LVEF ≥50% and are not captured by an exclusively LVEF-based approach. Therefore, we tested whether integration of an early disease stage with preserved LVEF (i.e. frequent PVCs/NSVT or LV LGE with LVEF ≥50%) would improve upon the LVEF-only based classification. Time event analysis demonstrates that this approach further stratifies an early disease stage with substantial residual risk compared to patients not meeting our proposed diagnostic criteria for DSP cardiomyopathy (**Figure 3**, **Table 4**). Greater LGE extent was only borderline associated in the univariate analysis (>4 segments, HR 1.38, p=0.05) and not additive to the multivariable analysis beyond PVCs/NSVT and LVEF (Supplemental Table 2). Therefore, LGE was used for identification of early disease, but was not directly used for determination of intermediate or advanced disease.

**Figure 3.**
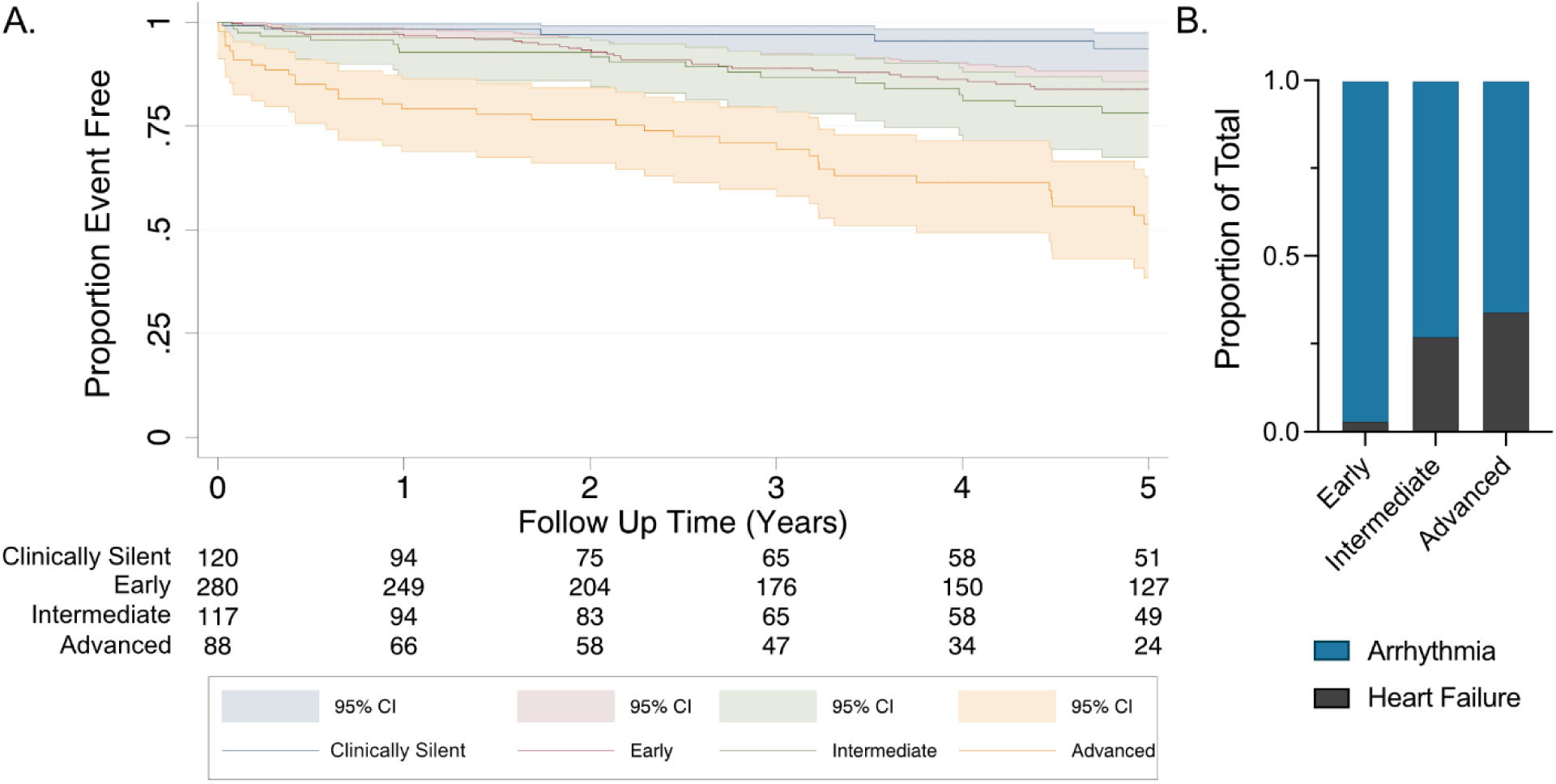
Adverse events by disease stage over 5 years of follow up. **A.** Composite adverse events (major arrhythmia and major heart failure) are shown up to 5 years of follow-up after baseline comprehensive diagnostic testing. **B.** Adverse event proportions are shown by ventricular arrhythmia or heart failure subtype for each disease stage. Corresponding Cox regression statistics are shown in Table 4.

**Table 4:** Cox regression analysis for adverse event by disease stage.

| Variable | HR | C.I. | p-value |
| --- | --- | --- | --- |
| Early Stage | 2.72 | 1.07-6.95 | 0.036 |
| Intermediate Stage | 3.70 | 1.38-9.91 | 0.009 |
| Advanced Stage | 10.3 | 4.01-26.2 | <0.001 |

Increased event rates in the intermediate and advanced stages were driven both by major arrhythmias (Supplemental Figure 3) and major heart failure events (Supplemental Figure 4; event types are shown in Supplemental Table 3). Notably, this approach also enabled identification of a low risk group – only five events were observed over 5-year follow-up in the phenotype negative group as defined by our proposed diagnostic criteria (N=120). In contrast, the event rate was high in the advanced stage group (n=88) with 38 events by 5 years (43%) (**Figure 3**). Taken together, these data show that disease staging incorporating DSP-specific features with LVEF-based classifications improves identification of risk for both heart failure and ventricular arrhythmia events.

### LV LGE Patterns Across Disease Stages

We next assessed LGE extent and distribution across disease stages using the 36-segment model that captures both regional and muscle layer involvement (see Methods and **Figure 4**). LGE localized to the subepicardial muscle layer in most patients who exhibited LGE (N=354/409, 87%). The most commonly affected segments were the subepicardial inferior or subepicardial lateral segments at the base of the ventricle (N=205/409, 50%) whereas relative sparing of the apical regions was observed (**Figure 4**). When stratified by the disease stages, patients with early disease demonstrated substantial LGE (mean of 4 regions; distribution shown in **Figure 4**). The extent of LGE was increased in patients with intermediate (mean of 5 regions, p<0.001) or advanced disease stage (mean of 5 regions, p=0.008). Supplemental Table 4). In a large number of patients, LGE was present in a circumferential ring-like pattern, and this pattern was more common in later disease stages (48% in early stage, 67% in intermediate stage, and 65% in advanced stage; p<0.001 early vs advanced, Supplemental Table 3). Additionally, we assessed for the presence of acute myocardial injury episodes across disease stages and found no significant differences (Supplemental Table 5). LGE extent was also similar in patients with or without clinically overt acute myocardial injury episodes, with the exception of a modestly greater association in the early disease stage (Supplemental Table 5). Taken together, these findings demonstrate that subepicardial LV fibrosis is an early feature of DSP cardiomyopathy and that a larger fibrosis burden accompanies declining LV systolic function, whereas acute myocardial injury episodes occur across the disease spectrum.

**Figure 4.**
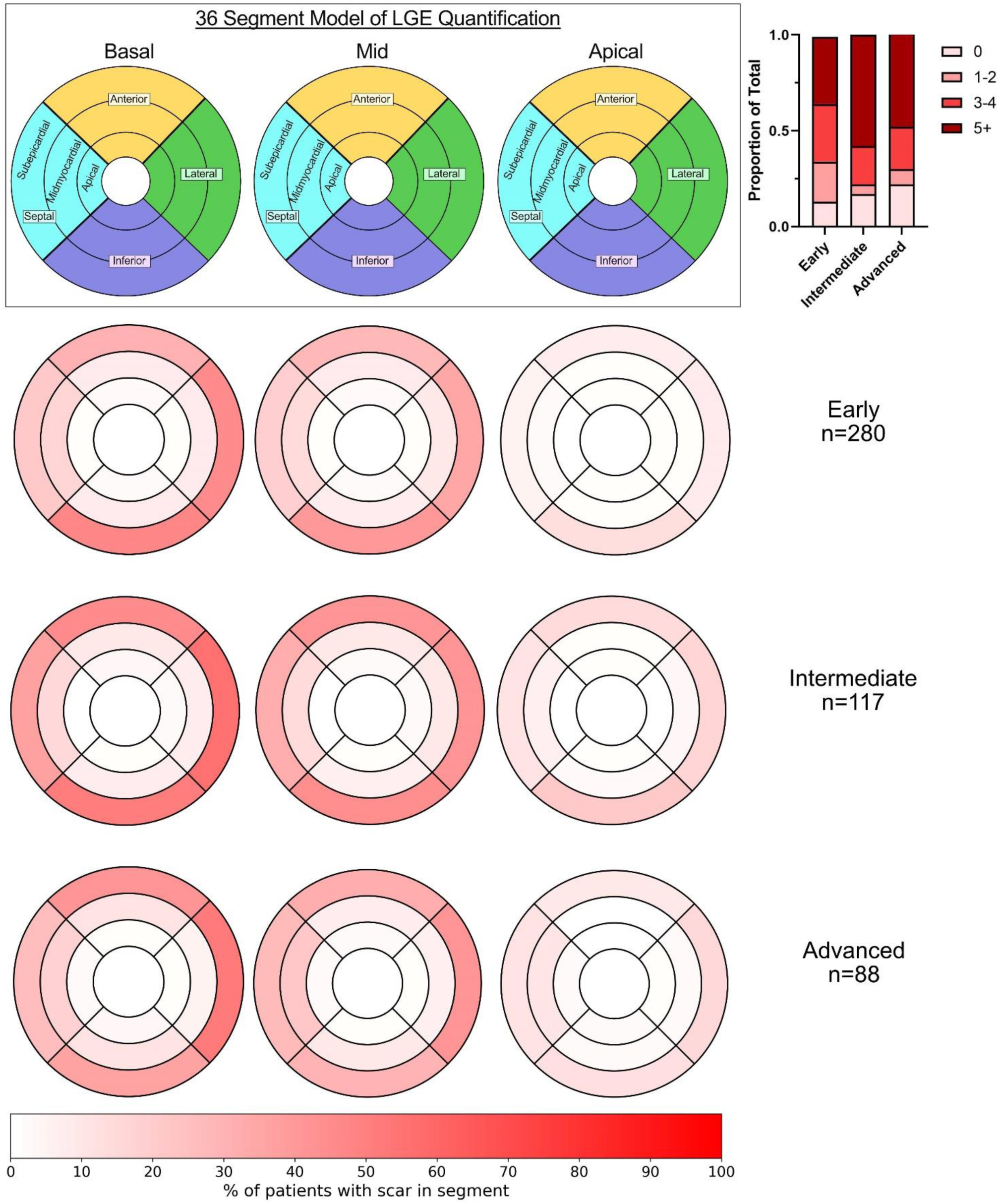
Late gadolinium distribution by disease stage and myocardial segment. A 36-segment model was used to capture presence/absence of late gadolinium enhancement (LGE) by region, ventricular level, and muscle layer (top row). The proportion of patients with LGE in each segment for each disease stage is shown by heat map (2^nd^ – 4^th^ rows). The relative proportion of patients with number of LGE segments by bin is shown for each disease stage (top right).

## Discussion

In this study, we have established a framework for gene-specific diagnostic criteria and disease staging for DSP cardiomyopathy. A strict requirement for inclusion of complete MRI and ambulatory rhythm monitoring at baseline enabled rigorous analysis of independent diagnostic contributions for key criteria. Moreover, inclusion of a large proportion of genotype positive family members allowed us to clearly define penetrance and evaluate the proposed staging scheme across the full disease spectrum. Integration of DSP-specific diagnostic and disease staging criteria with established ACC/AHA and ESC heart failure guidelines classifications improved risk assessment for both heart failure and major ventricular arrhythmia events and will facilitate straightforward incorporation into clinical practice.

### Diagnostic Criteria and Implications for Clinical Screening

There have been no genotype specific guidelines for DSP cardiomyopathy diagnosis to date, and criteria developed for ARVC diagnosis perform poorly.^1, 13, 14^ In the current study, we show that frequent PVCs (>500/24hr) or NSVT, reduction in LVEF (<50%), and subepicardial LGE on MRI are sensitive features of penetrant disease. Based on these findings, we propose a simple approach for highly sensitive diagnostic criteria that can readily be used in clinical practice (**Table 2**, **Figure 1**). Importantly, we demonstrated the clinical relevance of these diagnostic criteria by adverse event analysis, demonstrating that PVCs/NSVT and LVEF <50% strongly correlate with subsequent adverse events. Additionally, the presence of LGE exerts a clear additive effect with PVCs/NSVT on adverse event risk in patients with a preserved LVEF, enabling improved risk stratification in patients with early/mild disease. Moreover, the absence of any of the proposed diagnostic criteria identified a “clinically silent” group with low risk over the following 5 years. These diagnostic criteria will be particularly useful for guiding screening for family members who carry *DSP* pathogenic variants, particularly given the high clinical penetrance observed in this study. Our findings indicate that baseline screening including both ambulatory rhythm monitoring and MRI is essential given the sensitivity of both LGE and PVCs/NSVT for identifying clinical involvement.

### Disease Staging for Broad Risk Assessment Including Heart Failure

Beyond diagnostic criteria, we devised a disease staging algorithm for DSP cardiomyopathy that integrates with existing LVEF-based classifications. Prior studies of DSP cardiomyopathy have largely focused on ventricular arrhythmia risk.^1, 17^. In those studies, an LVEF <50% was identified as a threshold for sensitive identification of arrhythmia risk.^1, 17^ This finding was important, since a lower LVEF threshold, as used in typical DCM, was too permissive of arrhythmia risk. However, no prior studies have adequately investigated composite risk that includes outcomes related to progressive heart failure. Here, our objective was to evaluate the disease-driving parameters of LV fibrosis and declining systolic function to develop a disease staging approach reflective of fundamental disease progression in DSP cardiomyopathy. The availability of cardiac MRI on the entire cohort uniquely enabled a rigorous assessment of both of these parameters compared to prior studies.

We found that integration of LVEF categories supported in both the ACC/AHA and ESC guidelines improves identification of composite risk of both heart failure and ventricular arrhythmias. The greatest risk level was clearly evident in the LVEF <40% group – notably, this group exhibited a marked elevation in heart failure risk and also a substantial increase in major ventricular arrhythmia risk (2.9-fold and 2.3-fold increase in incidence rates compared to the “Intermediate” disease stage, respectively). Moreover, to create a comprehensive staging approach, we combined the LVEF classification with the diagnostic criteria to distinguish the “Early” disease stage (i.e., diagnostic criteria present but preserved LVEF). Designation of the early disease stage enables identification of individuals who should be followed more closely for progression but may not need aggressive interventions. This genotype-specific disease staging approach will also enable improved prognostication that will be clinically useful for discussion with patients in the clinic. We envision that the diagnostic and staging approach here can be used in tandem with the ventricular arrhythmia-focused risk calculator to yield an overall more comprehensive risk assessment.^17^ Finally, we anticipate that this approach will be critical for guiding clinical trial design for future gene-specific studies.

### Myocardial Fibrosis

Because we and others have previously observed a unique fibrosis pattern in DSP cardiomyopathy, we performed a semi-quantitative analysis of LGE localization using a 36-segment model that included localization by muscle layer. In this well-characterized cohort, we found that LV LGE is primarily subepicardial in DSP cardiomyopathy, confirming and extending results from prior studies. We also demonstrated that a high percentage of patients exhibit LGE even in the early disease stage, consistent with our previous report that LV LGE precedes overt systolic dysfunction.^1^ In early stage disease, we found strong interaction between LGE presence and frequent PVCs/NSVT driving ventricular arrhythmia risk, an important and clinically-relevant observation not captured in our prior analysis.^17^ Nevertheless, across the whole cohort, the semi-quantitative LGE burden did not strongly correlate with composite risk in our analyses, whereas frequent PVCs/NSVT and/or an LVEF <40% were strong multivariate risk predictors. Thus, this study indicates that LV fibrosis is a key feature driving pathogenesis of DSP cardiomyopathy and is key in identifying risk in early-stage disease with preserved LVEF. Additionally, LV fibrosis burden increases along with diminishment of LVEF consistent with a close relationship of these variables in disease progression. However, semiquantitative assessment of LGE burden, at least within the limitations of the present study, does not appear to incrementally add to ventricular arrhythmia risk assessment beyond LVEF and PVCs/NSVT for those with intermediate or advanced disease.

### Limitations

The findings of this study should be interpreted considering several limitations. Although many family members were included, patients in the DSP-ERADOS cohort were recruited at large academic centers – therefore referral bias may inflate estimations of both penetrance and adverse event risk. Nevertheless, our recruitment and analyses of non-proband, genotype-positive family members mitigate these limitations. MRI data was collected at individual sites, and central adjudication was not feasible for this project. This limitation may have led to variability in LGE assessment. Additionally, we did not analyze serial MRIs, which are seldomly available in patients after defibrillator placement. The current study also has the inherent limitations of retrospective cohort study. Future prospective registries may be useful to further refine these results and externally validate the diagnostic and staging scheme.

## Conclusion

In conclusion, we present a genotype specific approach for diagnosis and staging in DSP cardiomyopathy. We establish the sensitivity of three key clinical markers of disease and substantiate the clinical importance of these diagnostic criteria by analysis of adverse events. The proposed diagnosis and staging framework will aid in the clinical care of DSP patients across a broad range of cardiology providers and establish standardized terminology for future prospective research endeavors. The gene-centric approach used to establish criteria for diagnosis and staging in the current study will be widely applicable to other forms of inherited cardiomyopathy.

## Data Availability

Data supporting this manuscript will be made available upon reasonable request to the corresponding author

## Acknowledgements

Johns Hopkins myocarditis research is supported by the Matthew Poyner MVP Memorial Myocarditis Research Fund. The Zurich ARVC Program is supported by the Georg und Bertha Schwyzer-Winiker Foundation, Baugarten Foundation, USZ Foundation (Dr. Wild Grant), Swiss Heart Foundation and Swiss National Science Foundation.

## References

1. Smith ED, Lakdawala NK, Papoutsidakis N, Aubert G, Mazzanti A, McCanta AC, Agarwal PP, Arscott P, Dellefave-Castillo LM, Vorovich EE, Nutakki K, Wilsbacher LD, Priori SG, Jacoby DL, McNally EM, Helms AS. Desmoplakin Cardiomyopathy, a Fibrotic and Inflammatory Form of Cardiomyopathy Distinct From Typical Dilated or Arrhythmogenic Right Ventricular Cardiomyopathy. Circulation. 2020;141(23):1872–84. doi: 10.1161/CIRCULATIONAHA.119.044934.

2. Dalal D, Molin LH, Piccini J, Tichnell C, James C, Bomma C, Prakasa K, Towbin JA, Marcus FI, Spevak PJ, Bluemke DA, Abraham T, Russell SD, Calkins H, Judge DP. Clinical Features of Arrhythmogenic Right Ventricular Dysplasia/Cardiomyopathy Associated With Mutations in Plakophilin-2. Circulation. 2006-04-04;113(13). doi: 10.1161/CIRCULATIONAHA.105.568642.

3. Kumar S, Baldinger SH, Gandjbakhch E, Maury P, Sellal J-M, Androulakis AFA, Waintraub X, Charron P, Rollin A, Richard P, Stevenson WG, Macintyre CJ, Ho CY, Thompson T, Vohra JK, Kalman JM, Zeppenfeld K, Sacher F, Tedrow UB, Lakdawala NK. Long-Term Arrhythmic and Nonarrhythmic Outcomes of Lamin A/C Mutation Carriers. Journal of the American College of Cardiology. 2016/11/29;68(21). doi: 10.1016/j.jacc.2016.08.058.

4. Gigli M, Stolfo D, Graw S, Merlo M, Gregorio C, Chen SN, Ferro MD, Paldino A, Angelis GD, Brun F, Jirikowic J, Salcedo EE, Turja S, Fatkin D, Johnson R, Tintelen JPv, Riele AST, Wilde A, Lakdawala NK, Picard K, Miani D, Muser D, Severini GM, Calkins H, James CA, Murray B, Tichnell C, Parikh VN, Ashley EA, Reuter C, Song J, Judge D, McKenna WJ, Taylor MR, Sinagra G, Mestroni L. Phenotypic expression, natural history and risk stratification of cardiomyopathy caused by Filamin C truncating variants. Circulation. 2021 Sep 30;144(20). doi: 10.1161/CIRCULATIONAHA.121.053521.

5. Paldino A, Dal Ferro M, Stolfo D, Gandin I, Medo K, Graw S, Gigli M, Gagno G, Zaffalon D, Castrichini M, Masè M, Cannatà A, Brun F, Storm G, Severini GM, Lenarduzzi S, Girotto G, Gasparini P, Bortolotti F, Giacca M, Zacchigna S, Merlo M, Taylor MRG, Mestroni L, Sinagra G. Prognostic Prediction of Genotype vs Phenotype in Genetic Cardiomyopathies. Journal of the American College of Cardiology. 2022;80(21):1981–94. doi: 10.1016/J.JACC.2022.08.804.

6. Verstraelen TE, Lint FHMv, Bosman LP, Brouwer Rd, Proost VM, Abeln BGS, Taha K, Zwinderman AH, Dickhoff C, Oomen T, Schoonderwoerd BA, Kimman GP, Houweling AC, Gimeno-Blanes JR, Asselbergs FW, Zwaag PAvd, Boer RAd, Berg MPvd, Tintelen JPv, Wilde AAM. Prediction of ventricular arrhythmia in phospholamban p.Arg14del mutation carriers–reaching the frontiers of individual risk prediction. European Heart Journal. 2021 Jun 11;42(29). doi: 10.1093/eurheartj/ehab294.

7. Wang W, Murray B, Tichnell C, Gilotra NA, Zimmerman SL, Gasperetti A, Scheel P, Tandri H, Calkins H, James CA. Clinical characteristics and risk stratification of desmoplakin cardiomyopathy. Europace. 2021 Aug 5;24(2). doi: 10.1093/europace/euab183.

8. Bariani R, Cason M, Rigato I, Cipriani A, Celeghin R, De Gaspari M, Bueno Marinas M, Mattesi G, Pergola V, Rizzo S, Zorzi A, Giorgi B, Rampazzo A, Thiene G, Iliceto S, Perazzolo Marra M, Corrado D, Basso C, Pilichou K, Bauce B. Clinical profile and long-term follow-up of a cohort of patients with desmoplakin cardiomyopathy. Heart Rhythm. 2022;19(8):1315–24; PMCID: 35470109.

9. Gasperetti A, Carrick RT, Protonotarios A, Murray B, Laredo M, Schaaf Ivd, Lekanne RH, Syrris P, Cannie D, Tichnell C, Cappelletto C, Gigli M, Medo K, Saguner AM, Duru F, Gilotra NA, Zimmerman S, Hylind R, Abrams DJ, Lakdawala NK, Cadrin-Tourigny J, Targetti M, Olivotto I, Graziosi M, Cox M, Biagini E, Charron P, Casella M, Tondo C, Yazdani M, Ware JS, Prasad SK, Calò L, Smith ED, Helms AS, Hespe S, Ingles J, Tandri H, Ader F, Peretto G, Peters S, Horton A, Yao J, Dittmann S, Schulze-Bahr E, Qureshi M, Young K, Carruth ED, Haggerty C, Parikh VN, Taylor M, Mestroni L, Wilde A, Sinagra G, Merlo M, Gandjbakhch E, Tintelen JPv, Riele ASJMt, Elliott PM, Calkins H, James CA. Clinical features and outcomes in carriers of pathogenic desmoplakin variants. European Heart Journal. 2024 Sep 17;46(4). doi: 10.1093/eurheartj/ehae571.

10. Mazzarotto F, Tayal U, Buchan RJ, Midwinter W, Wilk A, Whiffin N, Govind R, Mazaika E, Marvao Ad, Dawes TJ, Felkin LE, Ahmad M, Theotokis PI, Edwards E, Ing AY, Thomson KL, Chan LL, Sim D, Baksi AJ, Pantazis A, Roberts AM, Watkins H, Funke B, O’Regan DP, Olivotto I, Barton PJ, Prasad SK, Cook SA, Ware JS, Walsh R. Reevaluating the Genetic Contribution of Monogenic Dilated Cardiomyopathy. Circulation. 2020 Jan 27;141(5). doi: 10.1161/CIRCULATIONAHA.119.037661.

11. Walsh R, Thomson KL, Ware JS, Funke BH, Woodley J, McGuire KJ, Mazzarotto F, Blair E, Seller A, Taylor JC, Minikel EV, Exome Aggregation C, MacArthur DG, Farrall M, Cook SA, Watkins H. Reassessment of Mendelian gene pathogenicity using 7,855 cardiomyopathy cases and 60,706 reference samples. Genet Med. 2016;17(10):90-.

12. Gasperetti A, Carrick R, Protonotarios A, Laredo M, Schaaf Ivd, Syrris P, Murray B, Tichnell C, Cappelletto C, Gigli M, Medo K, Crabtree P, Saguner AM, Duru F, Hylind R, Abrams D, Lakdawala NK, Massie C, Cadrin-Tourigny J, Targetti M, Olivotto I, Graziosi M, Cox M, Biagini E, Charron P, Casella M, Tondo C, Yazdani M, Ware JS, Prasad S, Calò L, Smith E, Helms A, Hespe S, Ingles J, Tandri H, Ader F, Mestroni L, Wilde A, Merlo M, Gandjbakhch E, Calkins H, Riele ASt, Tintelen JPv, Elliot P, James CA. Long-Term Arrhythmic Follow-Up and Risk Stratification of Patients With Desmoplakin-Associated Arrhythmogenic Right Ventricular Cardiomyopathy. JACC: Advances. 2024 Feb 2;3(3). doi: 10.1016/j.jacadv.2024.100832.

13. Corrado D, Zorzi A, Cipriani A, Bauce B, Bariani R, Beffagna G, Lazzari MD, Migliore F, Pilichou K, Rampazzo A, Rigato I, Rizzo S, Thiene G, Marra MP, Basso C. Evolving Diagnostic Criteria for Arrhythmogenic Cardiomyopathy. Journal of the American Heart Association: Cardiovascular and Cerebrovascular Disease. 2021 Sep 17;10(18). doi: 10.1161/JAHA.121.021987.

14. Graziano F, Zorzi A, Cipriani A, Lazzari MD, Bauce B, Rigato I, Brunetti G, Pilichou K, Basso C, Marra MP, Corrado D. The 2020 “Padua Criteria” for Diagnosis and Phenotype Characterization of Arrhythmogenic Cardiomyopathy in Clinical Practice. Journal of Clinical Medicine. 2022 Jan 5;11(1). doi: 10.3390/jcm11010279.

15. Heidenreich PA, Bozkurt B, Aguilar D, Allen LA, Byun JJ, Colvin MM, Deswal A, Drazner MH, Dunlay SM, Evers LR, Fang JC, Fedson SE, Fonarow GC, Hayek SS, Hernandez AF, Khazanie P, Kittleson MM, Lee CS, Link MS, Milano CA, Nnacheta LC, Sandhu AT, Stevenson LW, Vardeny O, Vest AR, Yancy CW. 2022 AHA/ACC/HFSA Guideline for the Management of Heart Failure: A Report of the American College of Cardiology/American Heart Association Joint Committee on Clinical Practice Guidelines. Journal of the American College of Cardiology. 2022;79(17):e263–e421. doi: 10.1016/j.jacc.2021.12.012.

16. McDonagh TA, Metra M, Adamo M, Gardner RS, Baumbach A, Böhm M, Burri H, Butler J, Čelutkienė J, Chioncel O, Cleland JGF, Coats AJS, Crespo-Leiro MG, Farmakis D, Gilard M, Heymans S, Hoes AW, Jaarsma T, Jankowska EA, Lainscak M, Lam CSP, Lyon AR, McMurray JJV, Mebazaa A, Mindham R, Muneretto C, Piepoli MF, Price S, Rosano GMC, Ruschitzka F, Skibelund AK. 2021 ESC Guidelines for the diagnosis and treatment of acute and chronic heart failure. European Journal of Heart Failure. 2022;24(1). doi: 10.1002/ejhf.2333.

17. Carrick RT, Gasperetti A, Protonotarios A, Murray B, Laredo M, Schaaf Ivd, Dooijes D, Syrris P, Cannie D, Tichnell C, Gilotra NA, Cappelletto C, Medo K, Saguner AM, Duru F, Hylind RJ, Abrams DJ, Lakdawala NK, Cadrin-Tourigny J, Targetti M, Olivotto I, Graziosi M, Cox M, Biagini E, Charron P, Compagnucci P, Casella M, Conte G, Tondo C, Yazdani M, Ware JS, Prasad SK, Calò L, Smith ED, Helms AS, Hespe S, Ingles J, Tandri H, Ader F, Peretto G, Peters S, Horton A, Yao J, Schulze-Bahr E, Dittman S, Carruth ED, Young K, Qureshi M, Haggerty C, Parikh VN, Taylor M, Mestroni L, Wilde A, Sinagra G, Merlo M, Gandjbakhch E, Tintelen JPv, Riele ASJMt, Elliott P, Calkins H, Wu KC, James CA. A novel tool for arrhythmic risk stratification in desmoplakin gene variant carriers. European Heart Journal. 2024 Jul 16;45(32). doi: 10.1093/eurheartj/ehae409.

18. Richards S, Aziz N, Bale S, Bick D, Das S, Gastier-Foster J, Grody WW, Hegde M, Lyon E, Spector E, Voelkerding K, Rehm HL. Standards and Guidelines for the Interpretation of Sequence Variants: A Joint Consensus Recommendation of the American College of Medical Genetics and Genomics and the Association for Molecular Pathology. Genetics in medicine : official journal of the American College of Medical Genetics. 2015 Mar 5;17(5). doi: 10.1038/gim.2015.30.

19. Van-Lint FHM, Murray B, Tichnell C, Zwart R, Amat N, Deprez RHL, Dittmann S, Stallmeyer B, Calkins H, Van-der-Smagt JJ, Wijngaard Avd, Dooijes D, van-der-Zwaag PA, Schulze-Bahr E, Judge DP, Jongbloed JDH, Tintelen JPv, James CA. Arrhythmogenic Right Ventricular Cardiomyopathy-Associated Desmosomal Variants Are Rarely De Novo. Circulation: Genomic and Precision Medicine. 2019;12(8). doi: 10.1161/CIRCGEN.119.002467.

20. Laredo M, Charpentier E, Soulez S, Nguyen V, Martino A, Calò L, Ader F, Hermida A, Fressart V, Charron P, Kachenoura N, Gandjbakhch E, Redheuil A. Imaging Features of Desmoplakin Arrhythmogenic Cardiomyopathy: A Comparative Cardiac Magnetic Resonance Study. Journal of Cardiovascular Magnetic Resonance. 2025. doi: 10.1016/j.jocmr.2025.101867.

